# Benchmarking of Germline Copy Number Variant Callers from Whole Genome Sequencing Data for Clinical Applications

**DOI:** 10.1101/2024.07.12.24310338

**Authors:** Francisco M. De La Vega, Sean A. Irvine, Pavana Anur, Kelly Potts, Lewis Kraft, Raul Torres, Peter Kang, Sean Truong, Yeonghun Lee, Shunhua Han, Vitor Onuchic, James Han

**Author notes:** Correspondence: Francisco M. De La Vega, Tempus AI, Inc., Stanford University School of Medicine.; James Han, Illumina, Inc.

## Abstract

Whole-genome sequencing (WGS) is increasingly favored over other genomic sequencing methods for clinical applications due to its comprehensive coverage and declining costs. WGS is particularly useful for the detection of copy number variants (CNVs), presumed to be more accurate than targeted sequencing assays such as WES or gene panels, because it can identify breakpoints in addition to changes in coverage depth. Recent advancements in bioinformatics tools, including those employing hardware acceleration and machine learning, have enhanced CNV detection. Although numerous benchmarking studies have been published, primarily focusing on open-source tools for short-read WGS CNV calling, systematic evaluations that encompass commercially available tools that meet the rigorous demands of clinical testing are still necessary. In clinical settings, where the confirmation of reported CNVs is often required, there is a higher priority on sensitivity over specificity/precision compared to research applications. Moreover, clinical gene panel reporting primarily concerns whether a CNV affects coding regions or, in some cases, promoters, rather than the precise detection of breakpoints. This study aims to benchmark the performance of various CNV detection tools tailored for clinical reporting from WGS using reference cell lines, providing insights critical for optimizing clinical diagnostics. Our results indicate that while different tools exhibit strengths in either sensitivity or precision and are better suited for certain classes and lengths of variants, few can deliver the balanced performance essential for clinical testing, where high sensitivity is imperative. Generally, callers demonstrate better performance for deletions than duplications, with the latter being poorly detected in events shorter than 5kb. We demonstrate that the DRAGEN™ v4.2 CNV caller, particularly with custom filters on its high sensitivity mode, offers a superior balance of sensitivity and precision compared to other available tools.

## Introduction

The clinical utility of whole-genome sequencing (WGS) is based on its ability to provide a more comprehensive and accurate identification of genetic variants, including SNVs/indels mitochondrial variants, and CNVs, all within a streamlined workflow.^1–5^ The cost of short-read WGS has declined over the past decade, driven by advancements in sequencing chemistries, increased density of sequencing reads per flow cell surface, and the scalability of sequencing instruments.^6,7^ The cost of short-read WGS is nearly equivalent to that of whole-exome sequencing (WES) while providing at least an 8% increase in diagnostic yield of individuals with genetic disease^7–10^ Consequently, clinical laboratories are eager to leverage the benefits of this technology and require bioinformatics tools that can deliver accurate variant calls.^11^

CNVs contribute significantly to genetic diversity and disease,^12^ yet they pose substantial detection challenges, especially in targeted sequencing assays.^13^ These assays, which have been the mainstay of clinical testing, typically detect variants based on subtle changes in sequencing coverage depth across exon footprints.^14^ Notably, the sensitivity for detecting single-exon deletions or duplications—a critical metric for clinical testing—has been reported to be around 50% at typical sequencing depths for WES (80-120X).^15^ WGS is set to supersede WES and gene panels in clinical settings due to its superior CNV/structural variant (SV) detection capabilities, faster turnaround times, and decreasing costs.^2,16^ Crucially, WGS facilitates the detection of entire CNVs, often spanning across intronic sequences not included in targeted panel sequencing, allowing the identification of the breakpoints of such variants, thus providing an additional layer of data beyond depth measurements to pinpoint these variants accurately.^17^

Despite the appearance that CNV detection from WGS is a resolved issue, evidence to substantiate this claim remains scant. Recent advancements in CNV callers have predominantly focused on the identification of these variants from targeted sequencing data.^14,18,19^ Moreover, emerging methods, including deep learning, promise enhanced accuracy, but broad benchmarking is lacking.^20,21^ Nevertheless, systematic evaluations of these tools to meet clinical standards are essential. Tools for short-read WGS CNV calling must be rigorously evaluated for clinical applications, where orthogonal confirmation of CNVs is typically performed,^22^ emphasizing the need for high sensitivity over specificity/precision compared to research applications.

In this study, we aimed to evaluate CNV calling tools designed for short-read, PCR-free WGS data. We used cell lines with known CNVs to assess their suitability for clinical gene-panel reporting from 50X WGS data. We compared the efficacy of several CNV callers using the HG002 reference cell line and a panel of 33 additional cell lines, all with documented CNVs in a selection of clinically relevant genes.

## Materials and Methods

### Cell Lines Utilized

For whole-genome level benchmarking, we utilized the cell line characterized by the Genome-in-a-Bottle Consortium: HG002. Additionally, we selected 25 cell lines from the Coriell Institute catalog that have reported CNVs (Table 1). These cell lines include CNVs that overlap genes in a panel comprising 89 hereditary cancer genes, 79 cardiometabolic disease genes, and 20 rare genetic disease genes (a total of 184 genes after removing redundancies; Supplementary Table 1).

**Table 1:** Detailed Metrics of CNV Detection by Caller. The table indicates the gene(s) overlapped by a CNV, chromosome (Chr), length of relevant events (based on our calls with DRAGEN), number of exons overlapped by CNV, and the type of event, either deletion (DEL) or duplication (DUP). * in the “Type” column indicates homozygous event. Some of these cell lines have multiple events and large complex rearrangements not listed by Coriell. We examined likely true positives, and when deemed confident, we added them to the truth set.

| Coriell ID | Gene(s) affected | Chr | Length (kb) | # of exons | Type |
| --- | --- | --- | --- | --- | --- |
| HG00343 | *CHEK2* | 22 | 5 | 2 | DEL |
| HG00634 | *PALB2* | 16 | 13 | 1 | DUP |
| HG03694 | *ATM* | 11 | 16 | 4 | DUP |
| NA02325 | *AXIN1-MEFV-PKD1-TSC2* | 16 | >3,000 | 145 | DUP |
| NA02325 | *LZTR1-SMARCB1-CHEK2-NF2* | 22 | >5,500 | 60 | DUP |
| NA03330 | *PARK2* | 6 | 198 | 1 | DUP |
| NA03330 | *BRCA2-N4BP2L1* | 13 | 3,174 | 26 | DUP |
| NA03330 | *SUCLA2* | 13 | 1,924 | 38 | DUP |
| NA03330 | *PARK2* | 6 | 198 | 1 | DUP |
| NA04372 | *GALC* | 14 | 32 | 7 | DEL |
| NA04517 | *GALC* | 14 | 32 | 7 | DEL* |
| NA04520 | *TSC2* | 16 | 89 | 35 | DEL |
| NA05117 | *DMD* | X | 165 | 2 | DEL |
| NA08618 | *ATM-DDX10* | 11 | >3,500 | 125 | DUP |
| NA10283 | *DMD* | X | 358 | 16 | DEL* |
| NA11661 | *GAA* | 17 | 1 | 1 | DEL |
| NA13434 | *PLP1* | X | 1 | 2 | DEL* |
| NA13480 | *ELN* | 7 | 1,304 | 33 | DUP |
| NA13480 | *JAK2* | 9 | 133 | 3 | DUP |
| NA14626 | *BRCA1* | 17 | 6 | 1 | DUP |
| NA18668 | *CFTR* | 7 | 21 | 2 | DEL |
| NA18949 | *BRCA1* | 17 | 6 | 2 | DEL |
| NA19401 | *TK2* | 16 | 6 | 1 | DEL |
| NA20381 | *CLN3* | 16 | 1 | 2 | DEL |
| NA21698 | *PARK2-PACRG* | 6 | >4,500 | 1 | DEL |
| NA21939 | *FBN1* | 15 | 6 | 4 | DEL |
| NA22208 | *PCCA* | 13 | 147 | 8 | DEL |
| NA23599 | *MECP2* | X | 15 | 2 | DEL |
| NA23710 | *CDKL5* | X | 8 | 2 | DEL |
| ND01039 | *PARK2* | 6 | 156 | 1 | DEL |

### Sample Preparation and Sequencing

PCR-free WGS libraries from the DNA of Coriell Institute’s cell lines were sequenced to a mean depth of 50X using paired-end 2×150bp reads on the Illumina NovaSeq 6000 system. The reads were mapped to the human reference genome GRCh37 using the DRAGEN Secondary Analysis Platform.^23^

### CNV Detection Tools Evaluated

We assessed multiple CNV calling tools for WGS including Delly (v1.6 - paired-end and split-read analysis),^24^ CNVnator (v0.4.1 - sequencing depth analysis),^25^ Lumpy (v0.2.13 - <u>paired-end and split-read analysis</u>),^26^ Parliament2 (ensemble of multiple CNV callers),^27^ Cue (cue.v2.pt model - Deep Learning from alignments),^21^ and the DRAGEN 4.2 CNV caller, which combines sequencing depth and split-read analyses (DRAGEN Secondary Analysis Platform, Illumina, Inc.).^23^ We utilized the DRAGEN CNV caller in two modalities: default parameters, and SV-supported high sensitivity mode (hereafter referred to as DRAGEN HS). All other tools used the default settings recommended by their respective developers and started from the same BAM alignments carried out with the DRAGEN multi-genome (graph) aligner.^23^

### Benchmarking Data Analysis

Given our clinical focus, we evaluated CNV calls based on their potential to disrupt protein structure. For benchmarking, we defined true positives as events that overlapped with coding exons of canonical transcripts and matched the dosage direction of the CNV truth set. Thus, we counted events intersecting an exon where the dosage direction matched the truth set as true positives. Events not meeting this condition were considered false positives. To avoid double counting, we adjusted for events spanning multiple exons. Sensitivity and precision were calculated according to the definitions published by the Global Alliance for Genomes and Health (GA4GH) Benchmarking team.^9^

For whole-genome level analysis, we used the Genome-in-a-Bottle SV truth set for HG002 v0.6 on GRCh37 (hs37d5) as our truth set.^28^ We identified CNVs in the truth set that overlapped with coding exons for the canonical transcripts of all human genes in GRCh37. The analysis was confined to events ranging from 500bp to 100kb. The original truth set contained 13 deletions and 4 duplications, overlapping 45 and 8 exons of GRCh37 canonical transcripts, respectively. To enhance our statistical analysis, we also simulated additional gene models on top of other variants in the truth set. The exon structures of the genes *BRCA1*, *BRCA2*, *CHEK2*, *PLP1*, and *GAA* were used as templates for constructing new genes. However, to avoid problematic regions of the genome, the construction of synthetic genes was limited to the high-confidence regions of HG002 as identified by the GIAB Consortium and specified in the file HG002_SVs_Tier1_v0.6.bed. The simulation was also restricted so that the synthetic genes would not overlap with any real human genes. In two cases, a small adjustment was made to the position of the synthetic gene to explicitly support some duplication events we had in the data (the number of verified duplications was small, and we needed to make the most of the ones we had). This simulation added 47 deletions and 6 duplications, overlapping 94 and 19 exons, respectively.

For the additional 33 cell lines, we used the CNV annotations described on the Coriell Institute website as the truth set. Since the reported alterations for cell lines in the Coriell catalog are sometimes incomplete, we further curated the truth set by visually inspecting the alignments of any putative false positives from the callers within our gene panel of interest. If the signal presented in the coverage graphs was deemed convincing, the variant was added to the truth set (see Table 1). We also included seven cell lines in the study (not listed in the table) that harbor CNV events requiring a targeted caller due to extensive paralogy (e.g., *CYP2D6*, *GBA*, and *PMS2*). In this study, we excluded calls across these genes from our evaluation and treated these cell lines as ‘true negatives’ for the assessment of the false positive rate per sample.

### Custom artifact filter for DRAGEN HS

Filtering for DRAGEN HS calls is designed to retain maximum sensitivity in genes of interest while discarding as many false positives as possible for the use case of gene-panel reporting out of WGS. First, we rejected any records smaller than 500 bases (a size under which DRAGEN HS emits numerous false positives) and larger than 10 Mbp (where we observed several recurrent breakpoint-based artifacts spanning centromeres or extending into telomeres, not relevant for our use case). Calls based solely on junction reads over 100 kbp were also discarded due to recurrent artifacts; true calls in this range would typically have depth support as well. We also excluded any calls overlapping centromeres and telomeres. Finally, calls that had a reciprocal overlap of ≥90% with recurrent artifacts identified across the cell lines were discarded. The resulting CNV calls product of using the DRAGEN HS method plus the custom filters will be hereafter referred as DRAGEN HS-F.

## Results

### Whole-genome benchmarking

We evaluated CNV callers at the genome level using WGS data from the cell line HG002, which has been extensively characterized and for which a truth set for CNVs and structural variants is available in GRCh37.^29^ This cell line is derived from a healthy individual and therefore does not harbor many CNV that disrupt gene function, which are the focus of our evaluation. The truth set contained 13 deletions and 4 duplications, overlapping 45 and 8 exons of GRCh37 canonical transcripts, respectively. To improve our statistical analysis, we also simulated gene models on top of other variants in the truth set to increase the number of overlaps evaluated. This simulation added 47 deletions and 6 duplications, overlapping 94 and 19 exons, respectively.

We included five commonly used open-source WGS CNV callers in our evaluation: Delly,^24^ CNVnator,^25^ Lumpy,^26^ and Parliament2 (an ensemble caller).^27^ Additionally, we evaluated two new WGS CNV callers: Cue, a deep-learning-based caller,^21^ and the DRAGEN 4.2 CNV caller, part of the hardware-accelerated DRAGEN Secondary Analysis Platform.^23^ This platform integrates the outputs of a sequencing depth-based CNV caller and a breakpoint-based caller into a unified call set. The DRAGEN caller can also be parametrized to operate in a high-sensitivity mode (HS), where sensitivity is gained at the expense of precision. We further evaluated the DRAGEN HS calls after filtering them through a custom process (referred as DRAGEN HS-F; cf. Methods).

We evaluated sensitivity and precision considering only events that overlap exons of both canonical and synthetic transcripts. The truth set included a total of 60 deletions and 10 duplications, each overlapping one or more exons. Our results (Figure 1) show that the maximum combined sensitivity (for deletions and duplications) achieved was 83% (DRAGEN HS), while the maximum precision was 76% (Cue). Delly exhibited high sensitivity (77%) but the lowest precision, and despite having the highest precision, Cue’s sensitivity was relatively low (33%). Lumpy exhibited very low sensitivity and precision in our tests. The DRAGEN caller in high sensitivity mode (DRAGEN HS) achieved the best sensitivity, albeit at a lower precision (30%). However, after applying a set of custom filters (DRAGEN HS-F), the precision improved substantially, with only a small decrease in sensitivity (75%), delivering the best balance between sensitivity and precision.

**Figure 1.**
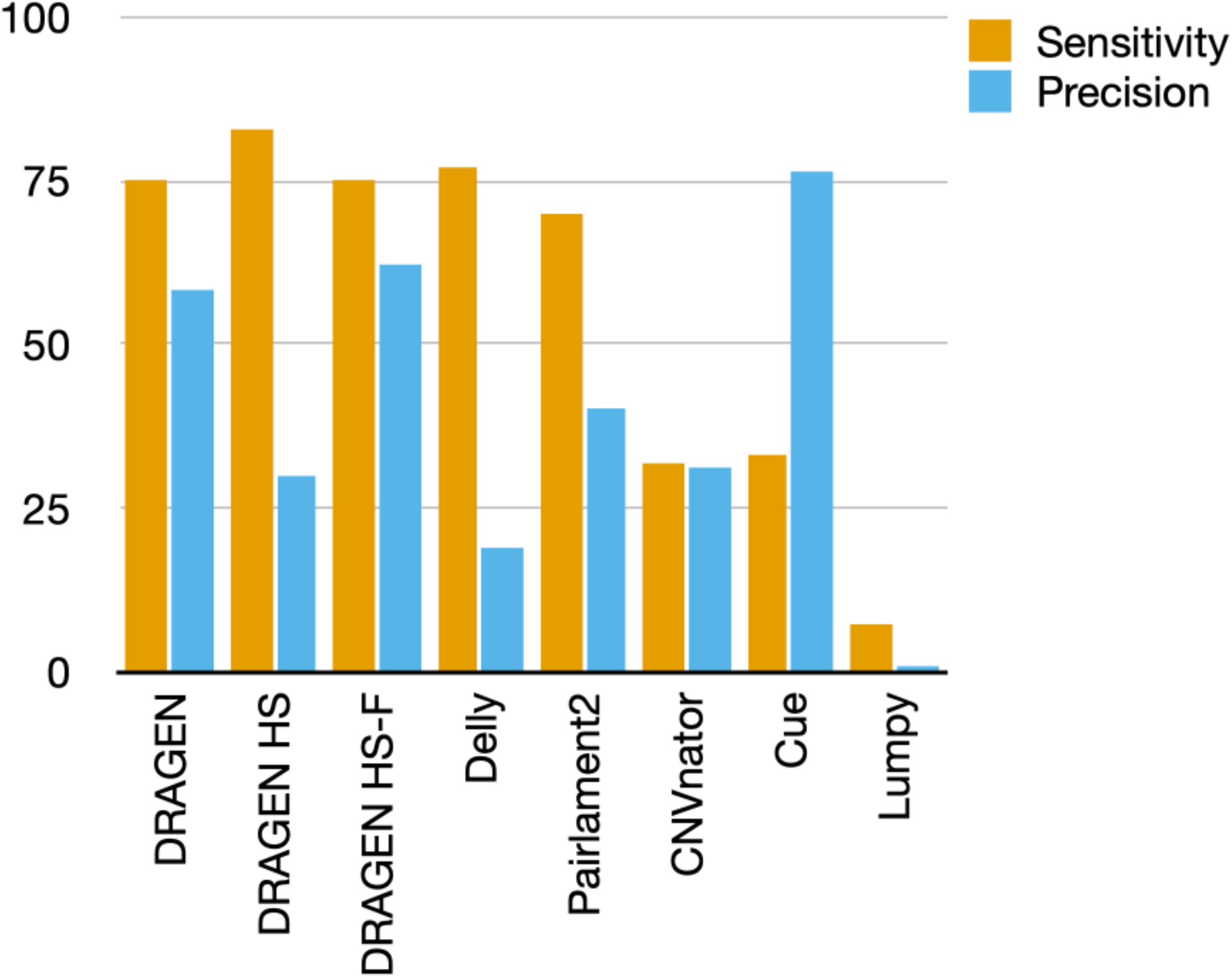
WGS Performance of the CNV/SV Callers Benchmarked. DRAGEN v4.2 CNV caller achieved the best balance of sensitivity and precision. The DRAGEN caller in high sensitivity mode (DRAGEN HS) had the highest sensitivity, albeit at a lower precision. On the other hand, Cue exhibited the best precision but had low sensitivity. We developed a set of custom filters for DRAGEN HS (cf. DRAGEN HS-F), which successfully improved precision with only a small reduction in sensitivity.

We then evaluated the performance of the callers, stratifying results by deletions and duplications (Figure 2). The performance for deletions followed a similar trend as described for the overall metrics (Figure 1). However, across all CNV callers, sensitivity was significantly lower for duplications. DRAGEN HS exhibited the highest sensitivity at 47%, while Cue achieved the best precision at 50%.

**Figure 2.**
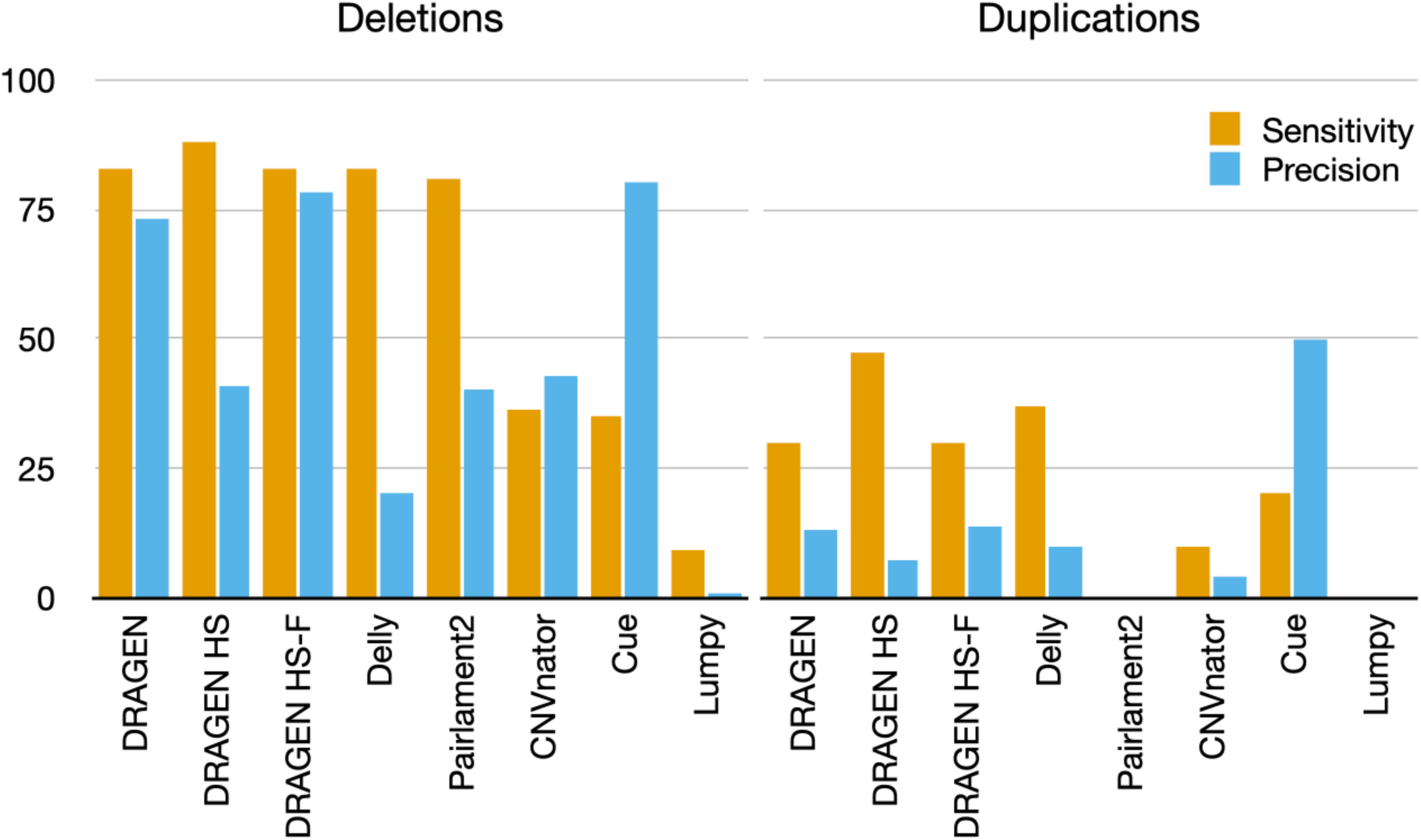
**Performance Exhibited by the Callers Stratified by CNV Type**: Deletions or Duplications. The performance for deletions follows a similar trend as described for overall metrics. Across all CNV callers, sensitivity was much lower for duplications, with DRAGEN HS exhibiting the highest sensitivity and Cue achieving the best precision.

Finally, we evaluated the combined performance (deletions and duplications) stratified by event length (Figure 3): 1-5 kb and >=5 kb (up to 100 kb). Generally, all callers exhibited lower sensitivity for events of 1-5 kb, with some unable to detect events in this range (e.g., CNVnator and Cue). Additionally, the sensitivity for duplications was lower across all callers, particularly for events of 1-5 kb. DRAGEN demonstrated the best precision for small duplications. Surprisingly, many callers showed very high sensitivity for duplications greater than 5 kb, although their precision remained low, except for Cue.

**Figure 3.**
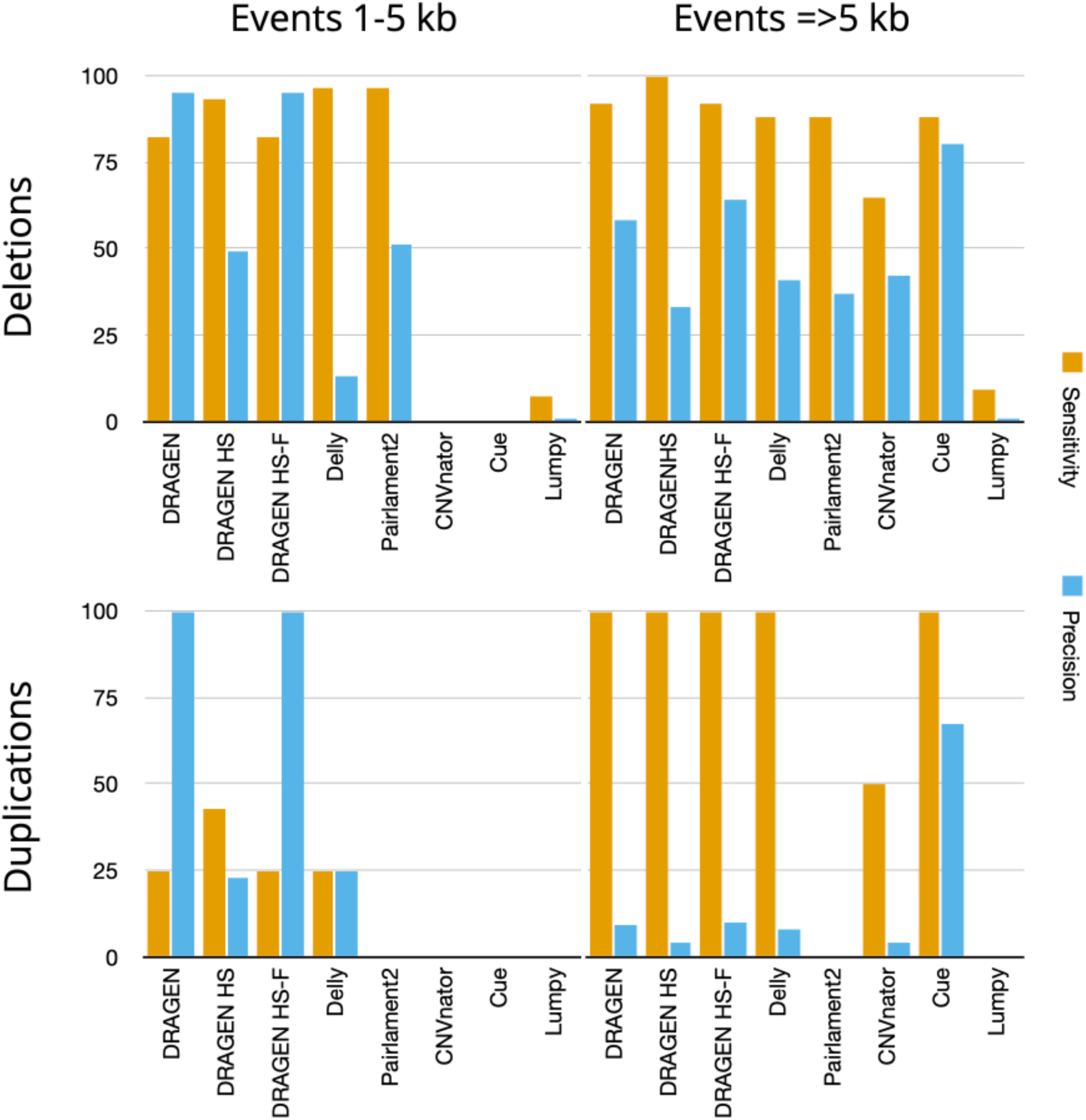
Performance by CNV Length. Performance is further stratified by event size, categorized as either 1-5kb or larger than 5kb. All callers exhibited lower sensitivity for smaller events, with some unable to detect events in this range. Sensitivity for duplications was particularly low for all callers, especially for events between 1-5kb.

### Benchmarking for virtual gene-panels from WGS data

We expanded our evaluation beyond the HG002 cell line by selecting cell lines from the Coriell Institute catalog that were annotated with CNVs in exons of a “virtual” panel of 184 genes commonly included in genetic testing for hereditary cancer, cardiovascular disease, and rare genetic disorders (Supplemental Table 1). Given that the rest of the genome in these cell lines has not been characterized, we limited our evaluation to the exons of the virtual panel. Furthermore, as we identified putative false positive calls during our evaluations, we examined the pattern of running depth coverage in windows along the genome to determine whether these false positives were, in fact, real events. This step was necessary because it is not uncommon for cell lines to harbor events not listed in the catalog metadata. Ultimately, we included several non-listed events in the truth set for evaluation.

In the 184 gene virtual panel, DRAGEN HS exhibited the highest sensitivity at 100%, while DRAGEN with standard settings achieved the highest precision at 77% (Figure 4). Notably, applying our custom filters to DRAGEN (HS-F) improved precision from 23% to 62% without reducing sensitivity. The other callers in the study demonstrated lower sensitivity and precision; however, surprisingly, Delly exhibited higher precision here than in the WGS evaluation. Given the promising results of DRAGEN HS in this setting, we further analyzed its performance stratified by CNV event size, either by the number of exons spanned or the length of the event (Table 2). The sensitivity remained at 100% across all strata, while precision for DRAGEN HS-F was 100% for events smaller than 1kb or overlapping one exon and decreased to 68% for events spanning five or more exons.

**Figure 4.**
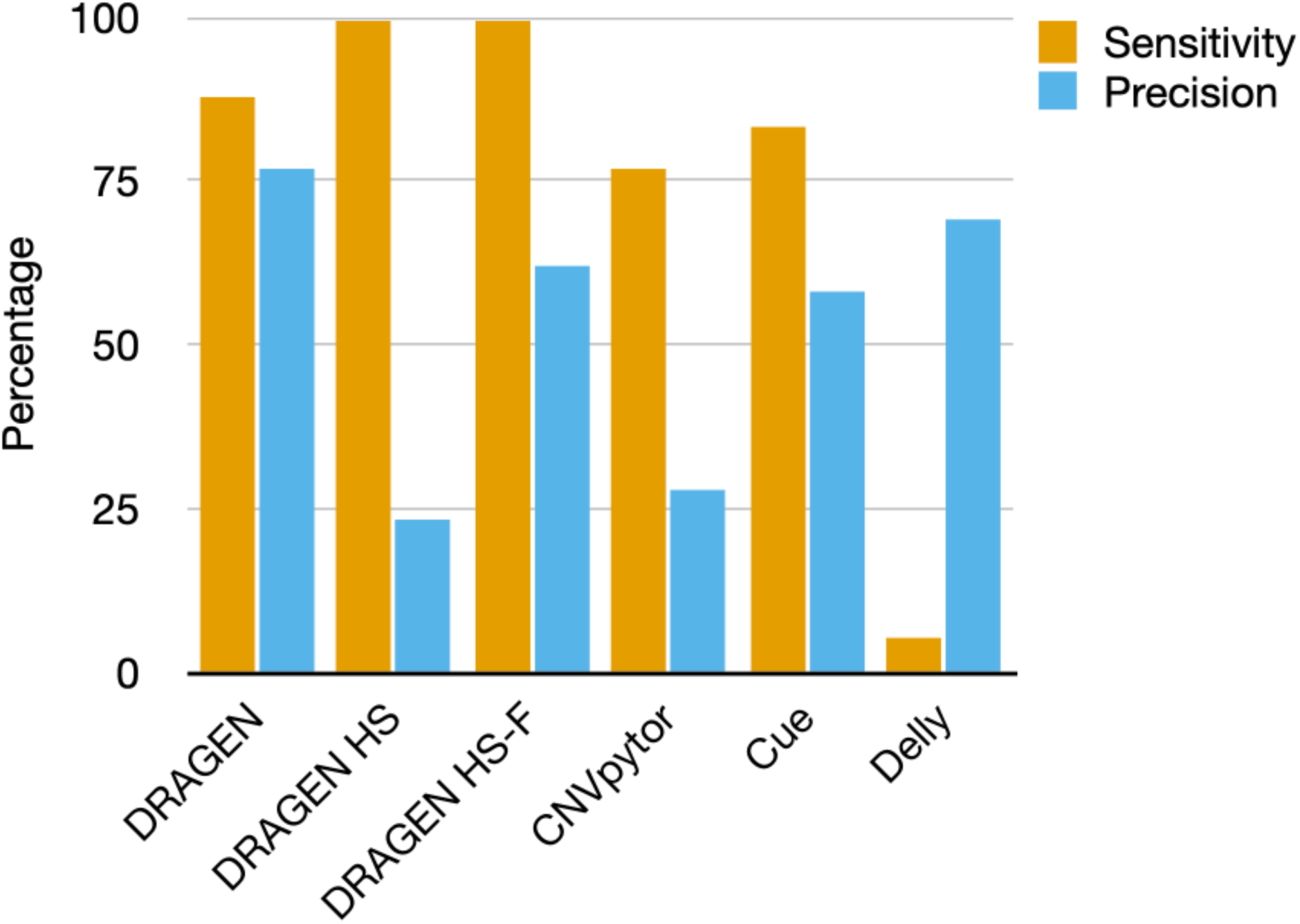
Gene-Panel Level Performance. This figure presents the overall performance of CNV callers benchmarked for the gene panel use case. The data show that the DRAGEN v4.2 CNV caller exhibited the best balance between sensitivity and precision. When set to high sensitivity mode (DRAGEN HS), it achieved the highest sensitivity, albeit at the cost of decreased precision. The use of custom filters (DRAGEN HS-F) successfully improved precision without sacrificing sensitivity. In contrast, the other callers in the study demonstrated lower sensitivity and precision.

**Table 2:** Detailed Metrics of CNV Detection for DRAGEN modalities.

| Exons spanned |  |  | CNV length |  |  |
| --- | --- | --- | --- | --- | --- |
| No. of exons | Precision HS (%) | Precision HS-F (%) | Length (kb) | Precision HS (%) | Precision HS-F (%) |
| 1 | 8 | 100 | 0.5 - 1 | 100 | 100 |
| 2 - 5 | 10 | 81 | 1 - 10 | 30 | 89 |
| > 5 | 1 | 68 | >10 | 2 | 74 |

To gain insights into failure modes, we examined the coverage patterns of genomic regions harboring examples of true and false positives from the DRAGEN HS-F caller. Figure 5A and 5B show examples of true positive deletions and duplications supported by both depth and junction breakpoints as a baseline. Figure 5C illustrates an example of a false positive, supported only by depth, that is caused by a mappability issue in regions paralogous to the *PMS2CL* pseudogene. Figure 5D depicts a false positive in *ACTN2*, which is actually a real deletion; however, the overextension of the 3’-end breakpoint results in a false positive exon overlap. In general, false positives were most commonly duplications ranging from 1-10kb in length.

**Figure 5.**
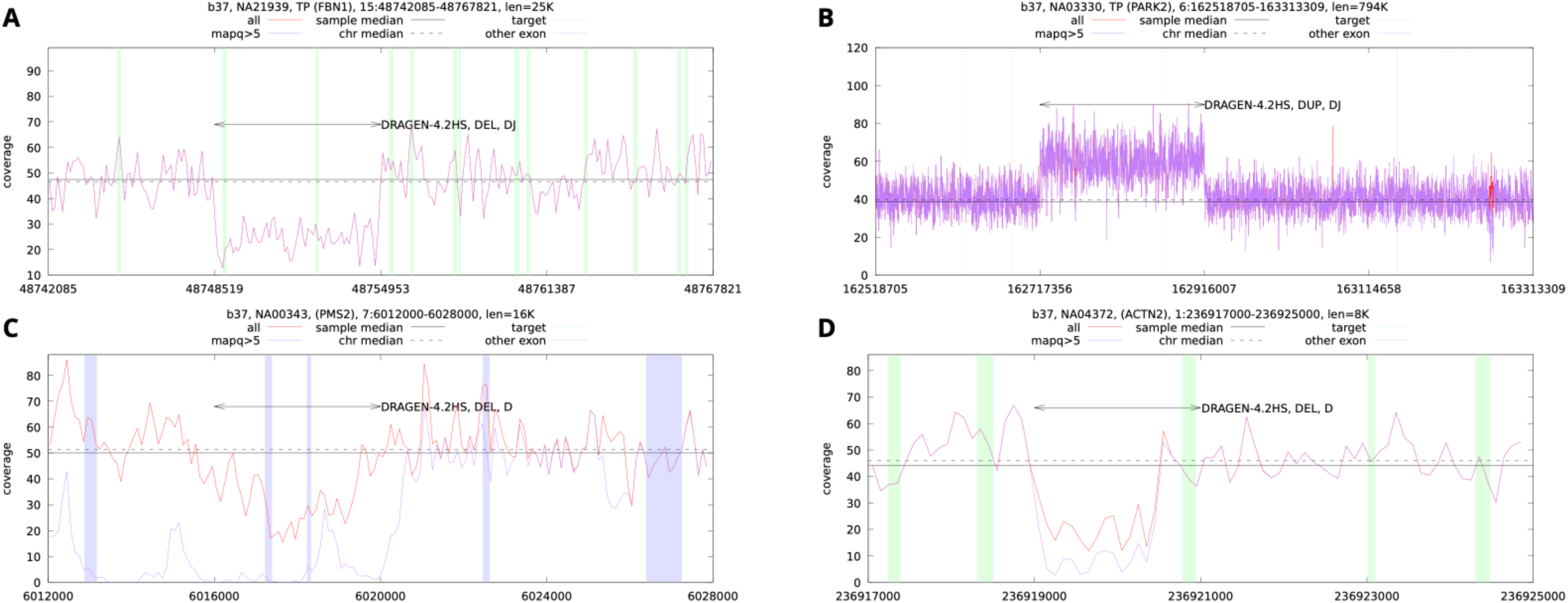
Examples of True Positive and False Positive CNV Calls for DRAGEN HS-F. Coverage graphs (100bp bins) show DRAGEN CNV calls marked with arrowheads. “D” indicates calls backed by depth analysis, and “J” denotes calls backed by junction reads (breakpoints). Shaded green vertical areas represent exons of the canonical transcripts of genes in the panel, while other regions are shown in blue. **Panel A** displays a true positive (TP) deletion (DEL) in *FBN1*; **Panel B** shows a TP duplication (DUP) in *PARK2*; **Panel C** illustrates a false positive (FP) deletion in the *PMS2* gene, caused by mappability issues in regions paralogous to the *PMS2CL* pseudogene (evidenced by a drop in MAPQ>5 coverage line); **Panel D** depicts a TP deletion in *ACTN2* where the overextended right breakpoint results in a FP exon overlap.

## Discussion

Given the relevance of CNVs in genetic diseases,^12^ the performance of CNV calling tools using NGS data can significantly impact the accuracy of clinical tests. Improving sensitivity for CNV detection in targeted sequencing panels—the workhorse of clinical testing to date—has been challenging, particularly for single-exon events.^15^ This difficulty arises from the limited data available to distinguish a diploid status from deletions or amplifications based solely on sequencing coverage depth, especially given the short length of typical exons.^14,30^ Some of this can be compensated for by increasing the overall sequencing depth (e.g., from ∼200-300X to >=500X) at the expense of increased assay cost. This situation is poised to improve with the decreasing costs of whole-genome sequencing (WGS) in recent years and its growing adoption in clinical testing.^7,8^ Unlike targeted gene panels or whole-exome sequencing (WES), which only capture and sequence coding exon regions, WGS provides data across the entire span of a CNV, often including non-coding regions and breakpoints.^17^ This additional information can be incorporated into CNV algorithms to improve the sensitivity of variant detection, even if the event only overlaps a single exon. Numerous bioinformatics tools have been developed over the past decade for this purpose.^14,18,19^ However, comprehensive benchmarking of these tools, focused on the goals of clinical genetic testing, is still lacking, particularly for those that include widely available commercial software used in top clinical labs.

We set out to assess and benchmark a set of commonly used WGS CNV callers that we were able to operate, including two recently released callers that implement innovative approaches: Cue, which uses a novel deep learning paradigm for structural variant identification,^21^ and DRAGEN 4.2 CNV caller, part of an FPGA-accelerated bioinformatics software suite that consolidates two independent approaches for CNV calling, namely, depth and breakpoint-based.^23^ In clinical genetic testing, the relevance of CNVs is established based on their ability to disrupt coding regions and impact the gene’s protein sequence. We, therefore, evaluated the callers based solely on how they overlap with coding regions, rather than attempting to assess the accuracy of their breakpoint identification or length. Importantly, we conducted evaluations for use cases where sensitivity needs to be very high (>99%), accepting lower precision because missing a clinically relevant variant in medical genetic testing is not acceptable.^22^ This lower precision is often managed by performing an orthogonal validation with a secondary technology (e.g. microarrays, RT-PCR, long-read sequencing) as part of the testing process to achieve a test-level precision of >99%.^22^ As long as the rate of CNV positive cases and false positives remains within single digits, this cost is amortized across all cases favorably. Thus, we prioritized sensitivity over precision in this evaluation.

As ground truth, we utilized data from a cell line used as a genomic standard by NIST and characterized as part of the Genome-in-a-Bottle consortium (HG002),^31^ developing a truth set of SVs and CNVs to use as a reference at the whole-genome level.^29^ Furthermore, we analyzed a series of cell lines previously described as harboring CNVs in a list of frequently tested genes. We analyzed this data focused on a virtual panel composed of genes that are typically tested for assays in hereditary cancer, cardiovascular disease, rare genetic disease panels.^2,5^ Performance was solely based on the coding regions of the genes in the aggregated panels encompassing 184 genes (Supplementary Table 1). Finally, we stratified our results by event type (deletion or duplication) and event size/length, to gain insights into the strengths and weaknesses of the different callers across these strata.

Our study provides a comprehensive evaluation of CNV detection tools in both whole-genome and gene-panel settings, including commercial tools available to clinical labs and that often are excluded in published benchmarks. It demonstrates the varying performance levels of different callers, particularly between deletion and duplication detection. DRAGEN HS emerged as the most sensitive tool, achieving 100% sensitivity, a crucial requirement for clinical applications where detecting every possible event can be vital. However, its precision was lower at initial settings, a challenge commonly faced in high-sensitivity callers. The ability to customize filters according to specific clinical needs further enhances its utility. The application of our custom filters (DRAGEN HS-F) significantly improved precision without sacrificing sensitivity, highlighting the potential of optimized bioinformatics pipelines to balance the trade-offs between sensitivity and precision effectively.

These findings underscore the importance of tailored computational strategies in enhancing the utility of genomic data for clinical diagnostics. However, the presence of false positives, especially in regions with complex genomic architectures like those near pseudogenes, implied the need for bespoke callers tailored to the specific genes of interest in difficult genes such as in the case of *CYP2D6*.^32^

## Conclusions

The study underscores the importance of continuous benchmarking and improvement of CNV detection tools to meet clinical demands. DRAGEN v4.2 HS-F, with its adjustable sensitivity and precision, emerges as a promising option for integrating WGS into clinical diagnostic pipelines. Continued improvement of CNV detection methods is needed to improve analytical accuracy and reduce the cost and dependence on orthogonal validation of WGS results.

## Data Availability

All data produced in the present study are available upon reasonable request to the authors

## Acknowledgements

We thank Vanessa Nepomuceno from the Tempus publications team for the copyediting review.

---

**Supplemental Table 1.** Gene list for evaluation of gene-panel mode.

|  |  |  |  |  |  |
| --- | --- | --- | --- | --- | --- |
| ABCC9 | CLN3 | HOXB13 | NF2 | RYR2 | TPM1 |
| ABCG5 | COL3A1 | JAK2 | NOTCH1 | SAMD9 | TRDN |
| ABCG8 | CRYAB | JPH2 | NTHL1 | SAMD9L | TSC1 |
| ACTA2 | CSRP3 | JUP | PALB2 | SCN5A | TSC2 |
| ACTC1 | CTNNA1 | KCNH2 | PARK2 | SDHA | TTN |
| ACTN2 | DDX41 | KCNJ2 | PCCA | SDHAF2 | TTR |
| AIP | DES | KCNQ1 | PCSK9 | SDHB | VCL |
| ALK | DICER1 | KIT | PDGFRA | SDHC | VHL |
| ANKRD26 | DMD | LAMP2 | PHOX2B | SDHD | WT1 |
| APC | DOLK | LDLR | PKD1 | SKI |  |
| APOB | DSC2 | LDLRAP1 | PKP2 | SLC2A10 |  |
| APOE | DSG2 | LIPA | PLN | SMAD3 |  |
| ATM | DSP | LMNA | PLP1 | SMAD4 |  |
| AXIN1 | EFEMP2 | LOX | PMS2 | SMARCA4 |  |
| AXIN2 | EGFR | LPA | PMS2CL | SMARCB1 |  |
| BAG3 | ELN | LZTR1 | POLD1 | SMARCE1 |  |
| BAP1 | EMD | MAX | POLE | SRP72 |  |
| BARD1 | EPCAM | MECP2 | POT1 | STK11 |  |
| BLM | ETV6 | MEFV | PRKAG2 | SUCLA2 |  |
| BMPR1A | FANCC | MEN1 | PRKAR1A | SUFU |  |
| BRCA1 | FANCM | MET | PRKG1 | TAFAZZIN |  |
| BRCA2 | FBN1 | MITF | PTCH1 | TCAP |  |
| BRIP1 | FBN2 | MLH1 | PTEN | TERC |  |
| CACNA1C | FH | MSH2 | PTPN11 | TERT |  |
| CASQ2 | FHL1 | MSH3 | RAD51C | TGFB2 |  |
| CAV3 | FLCN | MSH6 | RAD51D | TGFB3 |  |
| CDC73 | FLNA | MUTYH | RAF1 | TGFBR1 |  |
| CDH1 | FLNC | MYBPC3 | RB1 | TGFBR2 |  |
| CDK4 | GAA | MYH11 | RBM20 | TK2 |  |
| CDKL5 | GALC | MYH7 | RECQL | TMEM127 |  |
| CDKN1B | GALNT12 | MYL2 | RET | TMEM43 |  |
| CDKN2A | GATA2 | MYL3 | RIT1 | TNNC1 |  |
| CEBPA | GBA | MYLK | RNF43 | TNNI3 |  |
| CFTR | GLA | NBN | RPS20 | TNNT2 |  |
| CHEK2 | GREM1 | NF1 | RUNX1 | TP53 |  |

